# Knowledge, structural barriers and attitudes to HIV Rapid Start among New Jersey providers

**DOI:** 10.1101/2022.12.27.22283980

**Authors:** Debbie Y Mohammed, Russell Brewer, Jason Leider, Eugene Martin, Sunny Choe

## Abstract

**Background:** Rapid Start results in persons with HIV (PWH) initiating antiretroviral therapy (ART) in less than seven days. Benefits associated with Rapid Start include linkage to medical care and starting ART on the same day as diagnosis. These PWH were better retained in medical care and likely to achieve virologic suppression, in a shorter time, than those who did not have access to Rapid Start. Despite recommendations to initiate ART less than seven days after diagnosis, slow uptake of Rapid Start, in New Jersey were noted.

**Objective:** Identify knowledge, structural barriers and attitudes to Rapid Start, among New Jersey providers.

**Methods:** An electronic survey using Qualtrics consisting of 33 questions with the following domains: provider and practice characteristics (11), knowledge (1), structural barriers (9) and attitudes to diverse patient types was administered to New Jersey providers. The results were analyzed using descriptive statistics due to small numbers over strata. Approval to conduct this survey was obtained from the William Paterson University Institutional Review Board.

**Results:** The respondents were less than 55 years old (36/56, 64%), female (44/60, 73%), heterosexual (50/59, 85%), and nurse practitioners or physician assistants (41/59, 69%). Those who identified as internal medicine (9, 47%) or infectious disease (6, 60%) providers or worked in Ryan White (3, 30%) and non-Ryan White (6, 55%) practices correctly identified that integrase inhibitors had the lowest prevalence of transmitted resistance, when compared to those in private and other clinical settings. Newly diagnosed patients were referred for medical care in 37 (65%) of the medical sites. However, only providers from Ryan White (federally funded clinics for HIV patients) (64%) and non-Ryan White (73%) public sites reported co-located HIV testing sites. Seventy percent of medical sites reported that they offered same-day medical appointments. However, a lower proportion of private (62%), public Ryan White (55%), and other medical sites (36%) offered same-day appointments compared to public non-Ryan White sites (82%). Despite having staff available 40 hours per week (91%), only 55% of Ryan White sites offered extended office hours in the early morning, evenings, or on Saturdays. When compared to providers in public Ryan White sites, a higher proportions of providers in non-Ryan White sites were comfortable doing Rapid Start either on the day of or within one week of diagnosis, 72% and 82%, respectively, or starting ART before genotype results were available, 46% and 55%, respectively. Providers in public non-Ryan White sites were comfortable with Rapid Start for the following diverse groups of patients: with untreated mental illness (64%), engaging in unprotected sex (73%), with multiple partners (91%), actively using illicit drugs (91%), without health insurance (91%), homeless (100%), and with acute infection (82%).

**Conclusions:** Policy and administrative decisions are needed to eliminate structural barriers at the clinic level. Education on guideline recommendations and with diverse groups of patients will increase comfort with Rapid Start.

## Background

Diagnosis, timely linkage to medical care, and initiation of antiretroviral therapy (ART), for persons with HIV (PWH), are hampered by a historical, stepwise approach to care, despite advances in technology and available medications. Prior to 2009, traditional HIV testing was conducted using a blood specimen sent to the laboratory, and if the initial ELISA test was positive, a confirmatory Western blot was performed [1]. This process resulted in delays in the receipt of confirmed positive results and referral to medical care. Consequently, many PWH did not receive confirmatory results in a timely manner resulting in poor linkage rates [2-4].

New Jersey introduced an innovative statewide rapid HIV testing model starting in 2003 [5]. This included a rapid test, with laboratory testing for confirmation of preliminary positive test results. From 2009, a Rapid-Rapid test algorithm was instituted at all test sites to facilitate linkage to medical care on the same day as a presumptive positive result [6]. Despite this technological advance, delays to timely linkage to HIV care continued. The reasons for delayed treatment included waiting for the results of laboratory test results including HIV genotyping, prior to the medical appointment and initiation of ART. In New Jersey, from 2007 to 2015, 72% of PWH were linked to care within 90 days and 60% in less than 30 days, based on documented initial laboratory results in the surveillance database [7].

Rapid Start results in PWH initiating ART in less than seven days, and in many cases on the day of diagnosis [8]. Early studies in support of Rapid Start reported that PWH who initiated ART earlier were better retained in medical care and were more likely to achieve virologic suppression by twelve months [9-11]. In the pivotal study supporting Rapid Start in the United States, the authors reported that the majority of PWH started ART on the day of diagnosis, and the median time to viral suppression was 1.8 months [12]. Subsequently, other clinical sites reported timely linkage on the day of diagnosis, faster time and improved viral suppression after initiating Rapid Start [13, 14].

Since 2019, the WHO recommendations [8] were supported by the Panel on ART Guidelines for Adults and Adolescents with HIV [15]. Despite the compelling evidence, many providers remain hesitant to initiate Rapid Start for PWH in their clinical practice. The purpose of this survey was to identify barriers to Rapid Start, among diverse New Jersey providers, in 2021.

## Methods

The survey was developed in collaboration with the Gilead Sciences, Medical Science Liaison in New Jersey (5^th^ author). Two content experts reviewed the survey after development and provided comments prior to distribution to providers in New Jersey (2^nd^ and 3^rd^ authors).

Physician addresses and fax numbers were obtained from Doctor Databases [16]. The survey was distributed three times to 4,500 physicians, twice by mail and once time by fax, in one-month intervals (January to March 2021). In addition, the survey was distributed electronically to 5,000 nurse practitioners by the New Jersey State Nurses Association (January to March 2021).

Surveys were collected anonymously over a one-year period, January to December 2021, using the online survey tool, Qualtrics. A twenty dollar Amazon gift card was provided to respondents who voluntarily provided their names, using a separate survey. The response rate was less than 1%, with 69 completed surveys.

The survey instrument consisted of 33 questions with the following domains: provider and practice characteristics (11), knowledge (1), structural barriers (9) and attitudes to diverse patient types (12). A screening question requested respondents to identify if they were a prescribing provider, and only prescribers were allowed to continue with the survey. The knowledge question included identification of the ART drug class with the lowest prevalence of transmitted drug resistant mutations (TDRM) that included integrase inhibitors (INSTIs), non-nucleoside reverse transcriptase inhibitors (NNRTIs), nucleoside reverse transcriptase inhibitors (NRTIs), and protease inhibitors (PIs).

Respondents were asked to indicate on a 5-point Likert Scale how strongly they agreed that structural barriers were present in their clinic setting and their attitudes towards diverse Rapid Start for PWH. For the analysis, these responses were collapsed into two categories: agree (strongly agree, somewhat agree), and disagree (neutral, strongly disagree and somewhat disagree). Structural barriers included referral sites (community, co-located test sites (Yes, No), availability of a medical appointment on the same day as diagnosis, necessity for prior authorizations before receipt of ART, and demonstration of eligibility for federal funding prior to receiving a medical appointment. Respondents were asked if clinical sites had staffing for 40 hours weekly and whether there was availability of extended clinic hours in the early morning, evenings and/or Saturdays. The level of comfort with Rapid Start was evaluated by the following criteria: a) starting ART on the day of diagnosis, b) less than one week after diagnosis, and c) before genotype results were available. Comfort with Rapid Start for the following patients was evaluated: those with untreated mental illness, engaging in condomless sex, having multiple partners, with active drug use, uninsured, homeless, with an acute infection, and those who are mentally ready and judged likely to be adherent to medical care.

Provider characteristics included: age, sex, number of years working with PWH, and attendance at continuing education activities per year. Practice characteristics included type of setting (urban, suburban), type of practice (infectious disease [ID], internal medicine [IM], and other family and adolescent medicine]); number of providers (1-10, 11-50, more than 50), number of patients (1-100, 101-499, more than 499), and clinic type (Private, Public Ryan White, Public non-Ryan White, and Other [including correctional facilities, hospitals, Veteran Affairs, as well as other clinic sites]).

The results were analyzed with SAS 9.4, using descriptive statistics due to small numbers over strata. If there were fewer than five responses, then the results were not analyzed or reported. Approval for this survey was obtained from the William Paterson University Institutional Review Board.

## Results

A total of 69 medical providers responded to the survey. The majority of respondents were younger than 55 years of age (36, 55%), female (44, 73%), heterosexual (50, 85%), and either nurse practitioners or physician assistants (41, 69%) (Table 1). Twenty-seven (38%) respondents reported that they did not have any experience working with PWH. Continuing education activities were limited to the past year, with six (10%) reporting that they attended zero and twenty-five (43%) attending one to two. They worked mostly in urban settings (34, 58%), internal medicine, (27, 45%), in practices with more than 50 providers (46, 66%) and 12 (24%) reported that their practice served more than 499 patients. Clinical sites were private (21, 36%), with a similar proportion in public Ryan White and non-Ryan White sites and other.

**Table 1:** Provider and Clinic Characteristics, New Jersey, 2021^a^.

| **Characteristics** | **Total N (%)** |
| --- | --- |
| **Age (N=56)** |  |
| 25-34 | 15 (27) |
| 35-44 | 14 (25) |
| 45-54 | 7 (13) |
| >= 55 | 20 (35) |
| **Sex (N=60)** |  |
| Female | 44 (73) |
| **Sexual Orientation (N=59)** |  |
| Heterosexual | 50 (85) |
| **Nurse Practitioner/Physician Assistant (N=59)** | 41 (69) |
| **Number of Years, PWH Experience (N=69)** |  |
| 0 | 27 (38) |
| 1-20 | 23 (33) |
| 21-35 | 20 (29) |
| **Continuing Education per Year (N=59)** |  |
| 0 | 6 (10) |
| 1-2 | 25 (43) |
| 3 | 12 (20) |
| >3 | 16 (27) |
| **Setting (N=59)** |  |
| Suburban and Rural | 25 (42) |
| Urban | 34 (58) |
| **Type of Practice (N=60)** |  |
| Infectious Diseases | 10 (17) |
| Internal Medicine | 27 (45) |
| Other (Family and Adolescent Medicine) | 23 (38) |
| **Number of Providers (N=59)** |  |
| 1-10 | 13 (19) |
| 11-50 | 10 (15) |
| >50 | 46 (66) |
| **Number of Patients (N=50)** |  |
| 1-100 | 19 (38) |
| 101-499 | 19 (38) |
| >499 | 12 (24) |
| **Clinic-type (N=59)** |  |
| Private | 21 (36) |
| Public-Ryan White Funded | 11 (19) |
| Public-non-Ryan White Funded | 12 (20) |
| Other<sup>b</sup> | 15 (25) |

<sup>a</sup> Respondents did not answer all questions. <sup>b</sup> Other included correctional facilities, hospitals, Veteran Affairs, as well as other clinic sites.

### Knowledge

Knowledge of which ART drug class had the lowest prevalence of TDRM was similar by respondents’ age, sex, sexual orientation, number of years of experience caring for PWH, attendance at yearly continuing education activities, practice size, and setting (data not shown). When the type of practice was evaluated, providers who identified as internal medicine (9, 47%) or infectious disease (6, 60%) were able to correctly identify that INSTIs had the lowest prevalence of TDRMs in the United States (Fisher’s exact test, p < 0.05), (Figure 1a).

**Figure 1a:**
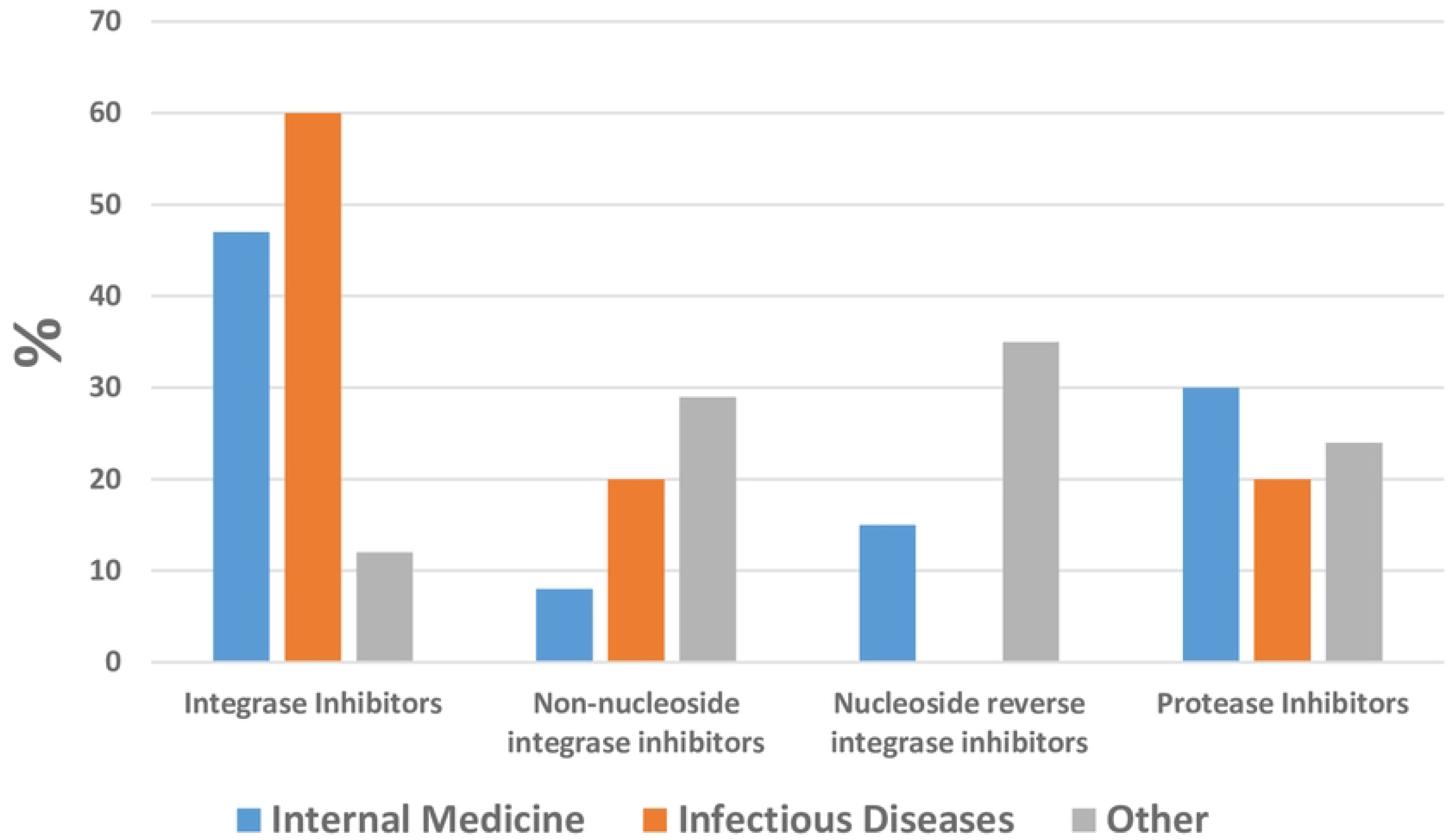
Knowledge of antiretroviral therapy with the lowest prevalence of transmitted resistance, by provider type. **Other: Family and Adolescent Medicine**

Providers in public settings, that is, Ryan White (3, 30%) and non-Ryan White (6, 55%) practices correctly identified that INSTIs had the lowest prevalence of transmitted resistance when compared to those in private and other clinical settings (Figure 1b).

**Figure 1b:**
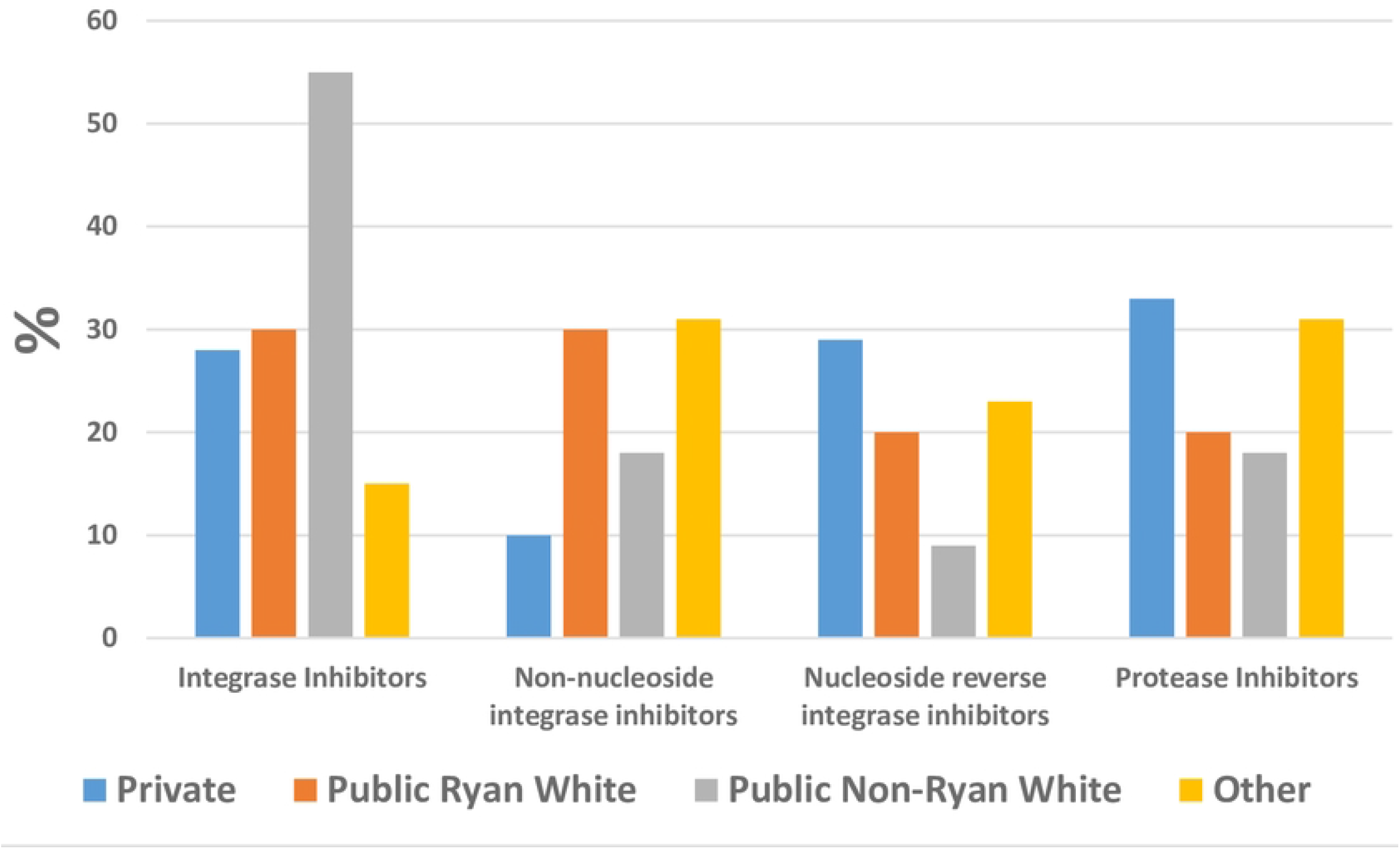
Knowledge of antiretroviral therapy with the lowest prevalence of transmitted resistance, by clinic type. **Other: Family and Adolescent Medicine**

### Structural Barriers to Rapid Start

Newly diagnosed patients were referred for medical care in 37 (65%) of the medical sites.

However, only providers from Ryan White (64%) and non-Ryan White (73%) public sites reported co-located HIV testing sites (Table 2a). Seventy percent of medical sites reported that they offered same-day medical appointments. However, a lower proportion of private (62%), public Ryan White (55%), and other medical sites (36%) offered same-day appointments compared to public non-Ryan White sites (82%).

**Table 2a:** Structural Barriers to Rapid Start, by Clinic Type, N=57.

| Characteristics | Private n=21 |  | Public Ryan-White n=11 |  | Public non-Ryan White n=11 |  | Other n=14 |  | Total n=57 |  |
| --- | --- | --- | --- | --- | --- | --- | --- | --- | --- | --- |
|  | Agree n(%) | Disagree n(%) | Agree n(%) | Disagree n(%) | Agree n(%) | Disagree n(%) | Agree n(%) | Disagree n(%) | Agree n(%) | Disagree n(%) |
| In my practice newly diagnosed patients |  |  |  |  |  |  |  |  |  |  |
| Are referred from the community | 14 (67) | 7 (33) | 7 (64) |  | 8 (73) |  | 8 (57) | 6 (43) | 37 (65) | 20 (35) |
| Are referred from collocated test sites | 9 (43) | 12 (57) | 8 (73) |  | 7 (64) |  | 7 (50) | 7 (50) | 31 (54) | 26 (46) |
| Can be scheduled for a same-day medical appointment | 13 (62) | 8 (38) | 6 (55) | 5 (45) | 9 (82) |  | 5 (36) | 9 (64) | 40 (70) | 17 (30) |
| Before receiving ART, prior authorizations are required by insurance companies | 8 (38) | 13 (62) |  | 7 (64) |  | 7 (64) |  | 10 (71) | 20 (35) | 37 (65) |
| Prior to receiving a medical appointment, PWH need to prove eligibility for federal funding | 7 (33) | 14 (67) |  | 7 (64) |  | 8 (73) |  | 10 (71) | 18 (32) | 39 (68) |
| The clinic site |  |  |  |  |  |  |  |  |  |  |
| Is staffed 40 hours/week | 11 (52) | 10 (48) | 10 (91) |  | 10 (91) |  | 8 (57) | 6 (43) | 39 (68) | 18 (32) |
| Has extended hours (early morning, late evening, Saturdays) | 8 (38) | 13 (62) | 6 (55) | 5 (45) |  | 7 (64) | 7 (50) | 7 (50) | 25 (44) | 32 (54) |
| I use pharmaceutical coupons for Rapid Start | 8 (38) | 13 (62) | 6 (60) |  | 7 (64) |  | 4 (31) | 9 (69) | 25 (45) | 30 (55)<sup>a</sup> |
| I use pharmaceutical drug samples for Rapid Start |  | 18 (85) | 4 (40) | 6 (60) | 5 (45) | 6 (55) | 6 (46) | 7 (54) | 18 (28) | 37 (72)<sup>a</sup> |

\*n=55; ART: Antiretroviral therapy

Access to ART or medical appointments was readily available as a high proportion of sites did not have to do prior authorizations (65%), and PWH did not have to demonstrate federal eligibility for medical services (68%). Despite having staff available 40 hours per week (91%), only 55% of Ryan White sites offered extended office hours in the early morning, evenings, or on Saturdays. Pharmaceutical coupons for Rapid Start were utilized at Ryan White (60%) and non-Ryan White (64%) sites; however 72% of sites did not have pharmaceutical samples readily available.

### Provider Attitudes to Rapid Start

Sixty-one percent of providers were comfortable with same-day Rapid Start, increasing to 77 percent less than one week after diagnosis. When compared to providers in public Ryan White sites, a higher proportion of providers in non-Ryan White sites were comfortable doing Rapid Start either on the day of or within one week of diagnosis, 72% and 82%, respectively, or starting ART before genotype results were available, 46% and 55%, respectively (Table 2b).

**Table 2b:**
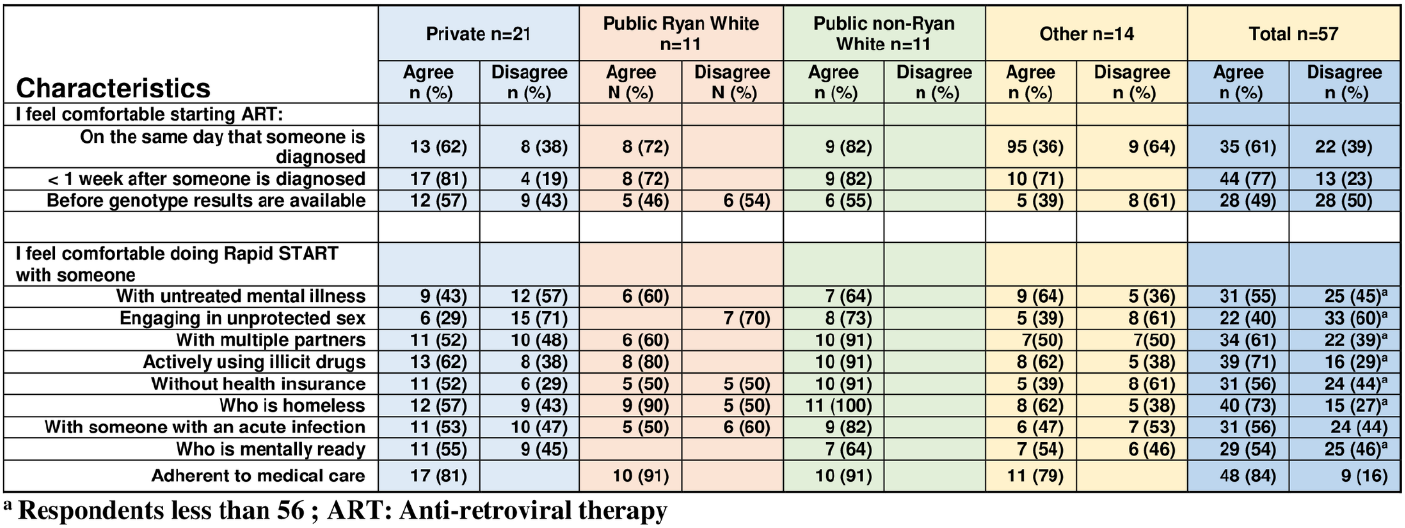
Provider Attitudes to Rapid Start, by Clinic Type, N= (56)

Providers in non-Ryan White sites were comfortable with Rapid Start for the following diverse groups of patients: with untreated mental illness (64%), engaging in unprotected sex (73%), with multiple partners (91%), actively using illicit drugs (91%), without health insurance (91%), homeless (100%), and with acute infection (82%).

## Discussion

Forty-four percent of respondents were not comfortable with Rapid Start for PWH without insurance, with higher proportions in Ryan White (50%) and other (61%) clinical sites. Access to Rapid Start on the day of or within one week of diagnosis requires coordination based on patients’ insurance, eligibility for charity care and federally funded programs. PWH without insurance may be eligible to apply for charity care or Ryan White funding, requiring documentation of residence and income. In an effort to support Rapid Start, for PWH without insurance, the Health Resources and Services Administration [17] emphasized that care and treatment services may be provided before eligibility is documented. Private sites will need to develop collaborations with publicly funded sites so that PWH can access medical care, including ART, on the same day or less than seven days after diagnosis.

Approximately one-third of the clinical sites reported that they completed prior authorization for ART. PWH with Medicaid as well as other insurance types may require prior authorizations when initiating ART. Many clinical sites may have staff that provide this support to providers and PWH. Access to ART can be facilitated using pharmaceutical coupons for copays, or samples kept on site. However, these strategies were not commonly employed by the respondents to this survey. Another recommended strategy to access ART, is the use of starter packs, until prior authorizations, copays and formulary issues are addressed [18].

Respondents were not comfortable with Rapid Start for PWH who were engaged in condomless sex (60%), had multiple partners (39%) or had an acute infection (44%), despite understanding that this strategy was effective in preventing transmission of HIV (93%). Support of Rapid Start for PWH engaged in condomless sex has been reported in two previous studies among heterosexual and gay couples. Among serodiscordant couples in which the HIV-positive partner was virologically suppressed on ART, and who reported condomless sex, there have been no documented cases of within-couple HIV transmission [19, 20]. The benefits of Rapid Start in acute infection are similar to persons with a chronic infection, including preventing transmission through rapid virologic suppression [21]. Rapid Start for PWH with acute infection can help maintain a healthy immune system; prior studies have demonstrated that earlier ART resulted in higher residual IFN-γ+ HIV-specific CD4 cells [22, 23]. More recently, acceptance of Rapid Start in acute infection was demonstrated, with over two-thirds (69%) of participants initiating ART within 7 days of diagnosis and 88% achieving viral suppression by week 48 [21].

Knowledge of the lowest prevalence of TDRMs in the United States, is key to Rapid Start, prior to the availability of genotype results. Multiple previous studies have reported on the prevalence of TDRM in the United States, including specific drug classes. Between 2009 and 2013, among patients enrolled in the Strategic Timing of Antiretroviral Treatment (START) study, the overall prevalence of TDRM was 2% for INSTIs, 9% for PIs, 13% for NRTIs, and 15% for NNRTIs [24]. Another study using surveillance data from 2014-2018 reported the prevalence of TDRM was 0.8% for INSTIs, 4.2% for PIs, 6.9% for NRTIs, and 12.0% for NNRTIs [25]. Current guidelines support starting an INSTI-containing regimen for newly diagnosed patients in the context of low TDRM [15]. Providers at all clinical sites in New Jersey, regardless of their practice or clinic type may benefit from knowledge of INSTIs as having the lowest prevalence of TDRM, thereby increasing their comfort with Rapid Start before HIV genotyping test results are available.

Improved linkage to medical care demonstrated by a visit with a prescribing provider is key to accessing Rapid Start. Same day medical appointments were not likely to be available in 30 percent of medical sites, despite having available staff for at least 40 hours/week, with some sites having early morning, evening, and weekend hours. The Rapid Entry and ART in Clinic for HIV (REACH) program reported that the median time to the first medical visit decreased from 17 to 5 days with viral suppression decreasing from 77 to 57 days [13]. Similarly, a previous study in New Jersey reported that a same day medical appointment increased viral suppression from 90% to 96% [14]. The study also reported a decreased median time to viral suppression after diagnosis, from 101 to 88 days, including a decreased time to receipt of ART, from 70 to 44 days, in a comparison of two time periods, 2013 to 2014, and 2016 to 2017, respectively. During 2013–2017, implementation of the Rapid ART Program Initiative for new Diagnoses (RAPID) in San Francisco, decreased the median time from diagnosis to viral suppression from 145 to 76 days, and the median time for ART initiation decreased from 28 to 1 days [26]. These data suggest that Rapid Start can be facilitated by increasing the availability of medical appointments with expanded hours for PWH on the day of diagnosis.

## Limitations

This study has the following limitations. Firstly, the study is limited to New Jersey providers. Secondly, the response rate was very low, limiting the generalizability to all New Jersey providers. The initial plan was to recruit respondents from large in-person conferences, but with limited in-person meetings from 2020-2021, due to the COVID-19 pandemic, this was not possible. Thirdly, the small sample size limited the ability to perform inferential statistics. Fourthly, self-report of the structural barriers and attitudes to Rapid Start, may be subject to social desirability bias. Lastly, 38% of the responders did not provide medical care for PWH.

## Conclusions

In summary, this survey identified gaps in knowledge related to TDRMs, clinic level structural barriers, and comfort with specific patient types that may impede Rapid Start.

Recommendations include policy decisions by funders and administrators to eliminate structural barriers at the clinic level, education on TDRMs, guideline recommendations for treatment initiation, including Rapid Start and that use of certain drug regimens (3-drug regimens containing 2nd generation INSTIs or boosted PIs) don’t lead to drug resistance, treatment as prevention for PWH, and management of diverse types of patients to increase comfort with Rapid Start.

## Data Availability

All relevant data are within the manuscript.

NA

